# Identifying Internal and External Shoulder Rotation Using a Kirigami-Based Shoulder Patch

**DOI:** 10.1101/2024.02.02.24302225

**Authors:** Amani A. Alkayyali, Conrad P. F. Cowan, Callum J. Owen, Emmanuel Giannas, Susann Wolfram, Ulrich Hansen, Alanson P. Sample, Roger J. H. Emery, Max Shtein, David B. Lipps

**Author notes:** Data availability statement*: The data that support the findings of this study are available from the corresponding author upon reasonable request. Author contributions statement*: Amani Alkayyali, Alanson Sample, and Max Shtein designed and manufactured the device used in data collection. Amani Alkayyali, Ulrich Hansen, Roger Emery, Max Shtein, and David Lipps developed the experimental protocol. Amani Alkayyali, Conrad Cowan, Callum Owen, and Emmanual Giannas conducted the experiments. Amani Alkayyali, Susann Wolfram, and David Lipps performed data analysis and summarization. Amani Alkayyali and David Lipps wrote the original manuscript. All authors contributed to data interpretation and participated in manuscript revisions. All authors have read and approved the final submitted manuscript. Ethics approval statement:* Ethical approval was granted by Imperial College London. Patient consent statement:* All participants reviewed their participation in the study with the research team and provided consent to participate in accordance with the ethics approval at Imperial College London. Corresponding author: David B. Lipps, Ph.D., School of Kinesiology, University of Michigan, 830 N. University Ave., SKB 1250, Ann Arbor, MI 48109.

## Abstract

Internal and external rotation of the shoulder is often challenging to quantify in the clinic. The current study evaluates a novel, engineered, wearable sensor system for improved internal and external shoulder rotation monitoring, and applies it in healthy individuals. Using the design principles of the Japanese art of *kirigami* (folding and cutting of paper to design 3D shapes), the sensor platform conforms to the shape of the shoulder with on-board strain gauges to measure movement. Our objective was to examine how well this *kirigami*-inspired shoulder patch could identify differences in shoulder kinematics between internal and external rotation as healthy individuals moved their humerus through specified movement patterns. Seventeen participants donned the wearable sensor on their right shoulder. Four strain gauges measured skin deformation patterns while participants moved their arm into internal or external rotation based on Codman’s paradox. One-dimensional statistical parametric mapping explored differences in strain voltage change of the strain gauges between internally-directed and externally-directed movements. The *kirigami* shoulder sensor, with its four on-board strain gauges, detected distinct differences in the movement pattern of participants who performed prescribed movements that resulted in either internal or external shoulder rotation. Three of the four strain gauges detected significant temporal differences between internal and external rotation (all p <0.047), particularly for the strain gauges placed distal or posterior to the acromion. These results are clinically significant, as they suggest a new class of wearable sensors conforming to the shoulder can measure differences in skin surface deformation corresponding to the underlying humerus rotation.

## INTRODUCTION

The prevalence of shoulder pain is between 18-26% of adults at any given time^1^ and negatively impacts activities of daily living (*e.g.*, showering, reaching for objects from a shelf, changing clothes). The presence of shoulder pain causes many individuals to adapt their shoulder movement patterns in adverse ways to complete functional tasks.^2–4^ The ability to monitor in detail the motion of the shoulder may allow for gauging and encouraging recovery, preventing further injury, and optimizing physical performance. However, the shoulder is the most complex joint in the human body, with the humerus capable of movement in six degrees of freedom, making it a challenge to measure the movement of the glenohumeral joint (e.g., humerus movement relative to the glenoid). Furthermore, restoration of internal and external rotation is particularly important for athletes participating in overhand throwing motions^5–7^ or patients recovering from total shoulder arthroplasty^8–10^.

It is challenging to accurately quantify the internal and external rotations of the shoulder in the clinic with current sensing modalities. Measuring these rotations with motion capture requires large, expensive experimental setups that are not feasible to replicate in a clinic. Inertial measurement units (IMUs) can be used in a clinical setting to quantify shoulder kinematics. This requires an accurate measure of orientation changes between sensors placed on the thorax and upper arm. However, the thorax sensor is difficult to secure and is the sensor location with the highest reported errors^11^. Measuring internal and external rotation is further complicated by Codman’s paradox^12^, which refers to specific movement patterns that allow an individual to move their shoulder in one dimension yet seem to achieve a change in two dimensions. Specifically, moving the humerus from a neutral position along the sagittal plane can achieve internal rotation without volitionally rotating the humerus. Similarly, moving the humerus from a neutral position along the coronal plane leads to external rotation without volitionally rotating the humerus. Conventional kinematic methods have not been successfully able to directly measure Codman’s paradox^13,14^, requiring new methods for quantifying internal and external shoulder rotation.

Traditional methods for evaluating and monitoring individuals with shoulder dysfunction in the clinic involve measuring joint range of motion (ROM), typically with goniometers^15^ or subjective questionnaires^16^. Higher fidelity data on shoulder range of motion can be collected in a laboratory setting with marker-based motion capture^16^ or wearable inertial measurement units (IMUs)^17^. Marker-based motion capture systems are large and expensive and ultimately not conducive to deploying for at-home monitoring of shoulder kinematics. While wearable IMU systems are more cost-effective and deployable, they can provide unreliable data at the shoulder, as there is a need for frequent recalibration to deal with signal drift and difficulty with the consistent, long-term placement of a sensor on the thorax to resolve humerothoracic angle calculations.

The current study evaluates a novel, engineered sensor system for improved internal and external shoulder rotation monitoring in healthy individuals. Using the design principles of the Japanese art of *kirigami* (folding and cutting of paper to design 3D shapes), the sensor platform starts out flat, permitting complex circuits to be defined on its surface using highly scaled industrial processes. At the same time, the cut pattern allows it to conform to the three-dimensional, curved shape of the shoulder, where on-board strain gauges can measure the extent of the substrate deformation and, therefore, joint movement.^18^ Our objective in the present study was to examine how well this *kirigami*-inspired shoulder sensor patch could identify differences in shoulder kinematics between internal and external rotation as healthy individuals moved their humerus through specified movement patterns corresponding to Codman’s paradox. To accomplish this, we created an untethered, wireless form factor, permitting free movement of the subject. This wearable sensor system, for the first time, allowed the quantitative measurement of movement patterns involved in Codman’s paradox. These results suggest utility for additional movement modalities, and a new class of wearable sensing at the shoulder that is currently unavailable with commercial systems and may be more conducive to at-home monitoring over existing platforms.

## MATERIALS AND METHODS

### Participants

Seventeen healthy individuals, all of whom are 18 or older, with no shoulder pathologies or upper extremity dysfunction, participated in this study [participant group data, mean (SD)] [11 male, age: 26 (4) years, height: 178 (7) cm, weight: 78 (9) kg, shoulder circumference: 45 (3) cm; 6 female, age: 23 (1) years, height: 170 (5) cm; weight: 61 (9) kg, shoulder circumference: 38 (2) cm]. Fifteen of 17 participants were right-hand dominant based on self-report.

Participants wore personal clothing of their choosing while exposing their right shoulder to make room for the shoulder patch hardware. All of the participants were recruited from Imperial College London, with ethical approval for this study granted by its institutional ethics committee.

### Shoulder Patch Hardware

Our team initially developed a *kirigami* patch that conforms to the shape of the shoulder to measure local changes in strain as an individual moves their shoulder^18^. This device has been subsequently upgraded to accommodate more strain gauges, and provide wireless data collection, powered by an on-board, rechargeable lithium-ion battery. The wearable comprises four main modules: 1) a custom-designed flexible printed circuit board (PCB) that houses four strain gauges and adheres to one’s skin with a skin-safe adhesive, 2) a custom-designed PCB with a Wheatstone bridge and operational amplifier circuitry to enhance the strain gauge signals, 3) a microcontroller unit (MCU) powered by a rechargeable lithium-ion battery, which samples the strain gauges at 96kHz and transports the measured data wirelessly *via* an onboard Bluetooth module to a host computer, and 4) a computer to display the and store data. The components of the device are shown in **Figure 1**.

**Figure 1:**
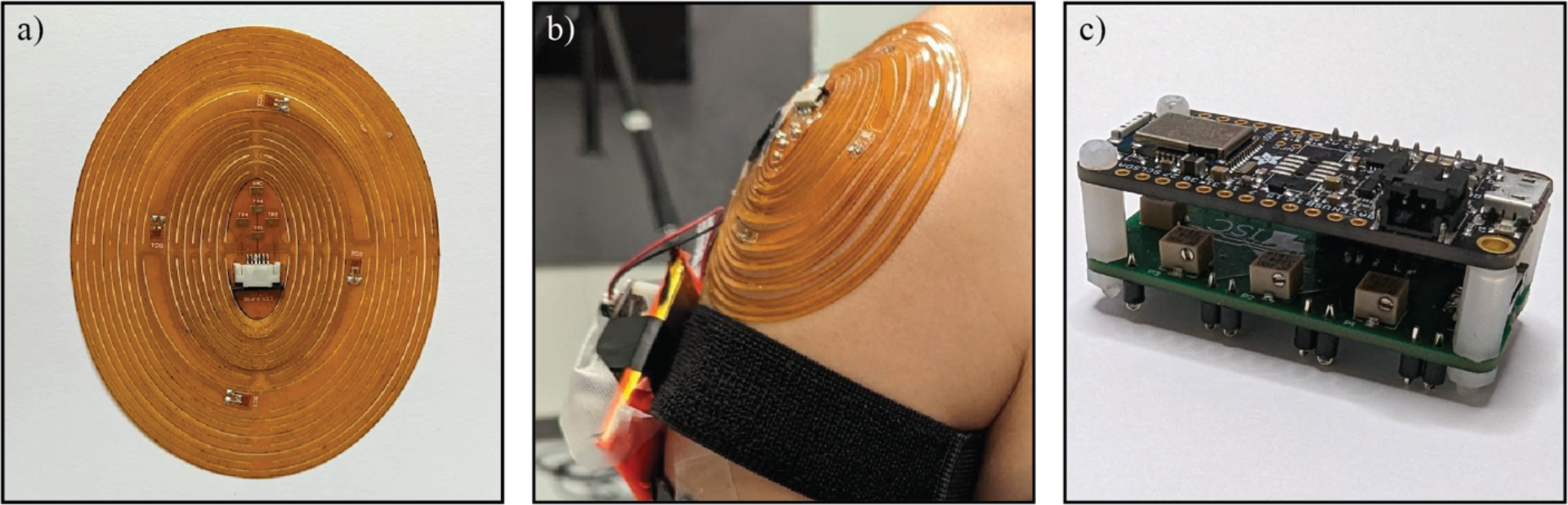
a) Top view of the kirigami-inspired shoulder patch lying flat with four strain gauges. b) Anterior view of the kirigami patch placed over the shoulder, with ribbon cabling from the patch to the electronics placed within an armband. c) The electronics contained within the armband, including a Wheatstone bridge board and Bluetooth module board.

The current *kirigami* patch includes four strain gauges (Omega uniaxial linear strain gauges, SGT-1/350-TY41) attached to the flexible PCB with double-sided adhesive. Signals from each of the four strain gauges (referred to as strain gauges 1-4) were recorded simultaneously. The placement of the patch on the shoulder and the corresponding strain gauge location are detailed below. The MCU was an Adafruit Feather nRF52 Bluefruit LE board, connected to the Wheatstone bridge with header pins. Data was collected using a custom Python script on a 2023 MacBook Pro, running MacOS Ventura.

Before attaching the patch, the skin of the shoulder was prepared using a skin exfoliant and alcohol wipes. If the participant had hair on their shoulder, it was shaved before using the exfoliant. While the patch incorporated an adhesive backing, the shoulder was sprayed with a skin adhesive (Cramer Tuf-Skin) before placing the patch to maximize adhesion. The same individual placed the *kirigami* patch on all participants to maintain consistency with particular care not to stretch or deform the patch when placing it on the shoulder. The patch was placed on the participant such that the center of the patch was placed atop the acromion, with azimuthal orientation ensuring that strain gauge 1 was located distal of the acromion on the humerus, strain gauge 2 was located anterior to the acromion, strain gauge 3 was located proximal to the acromion proximally, and strain gauge 4 was located posterior to the acromion (**Figure 1B)**.

### Protocol

All participants performed the same experimental protocol, in a single testing session lasting approximately one hour. All participants were tested on their right shoulder to maintain consistency in data collection and analysis. Moving their right arm according to Codman’s paradox^12^, each participant performed ten trials. Five trials resulted in internal rotation, and the other five resulted in external rotation. Trial order was randomized. Before completing the experimental trials, the research team demonstrated to each participant how to perform each task, performed the movement along with them, and provided verbal feedback to guide them to the correct position.

According to Codman’s paradox, a participant can move their arm in one rotation, yet appear to have moved it in two rotations. Mimicking the four movements described in Codman’s paradox, the experimental protocol required participants to begin in a neutral position, with their arms to their sides and palms facing forward. The participants then moved their right hand above their head as high as possible with their palm facing medially. Then, without moving the humerus, they bent their elbow at a right angle such that their forearm rested on the top of their head. For the external rotation task, with their elbow fixed at a right angle and without rotating their arm, the participants descended their humerus slowly to the side of their body in the coronal plane. For the internal rotation task with their elbow fixed at a right angle and without rotating their arm, the participants descended their humerus slowly in front of them in the sagittal plane. Lastly, each participant was instructed to return their arm from internal or external rotation to the neutral position. These motions are depicted in **Figure 2**.

**Figure 2:**
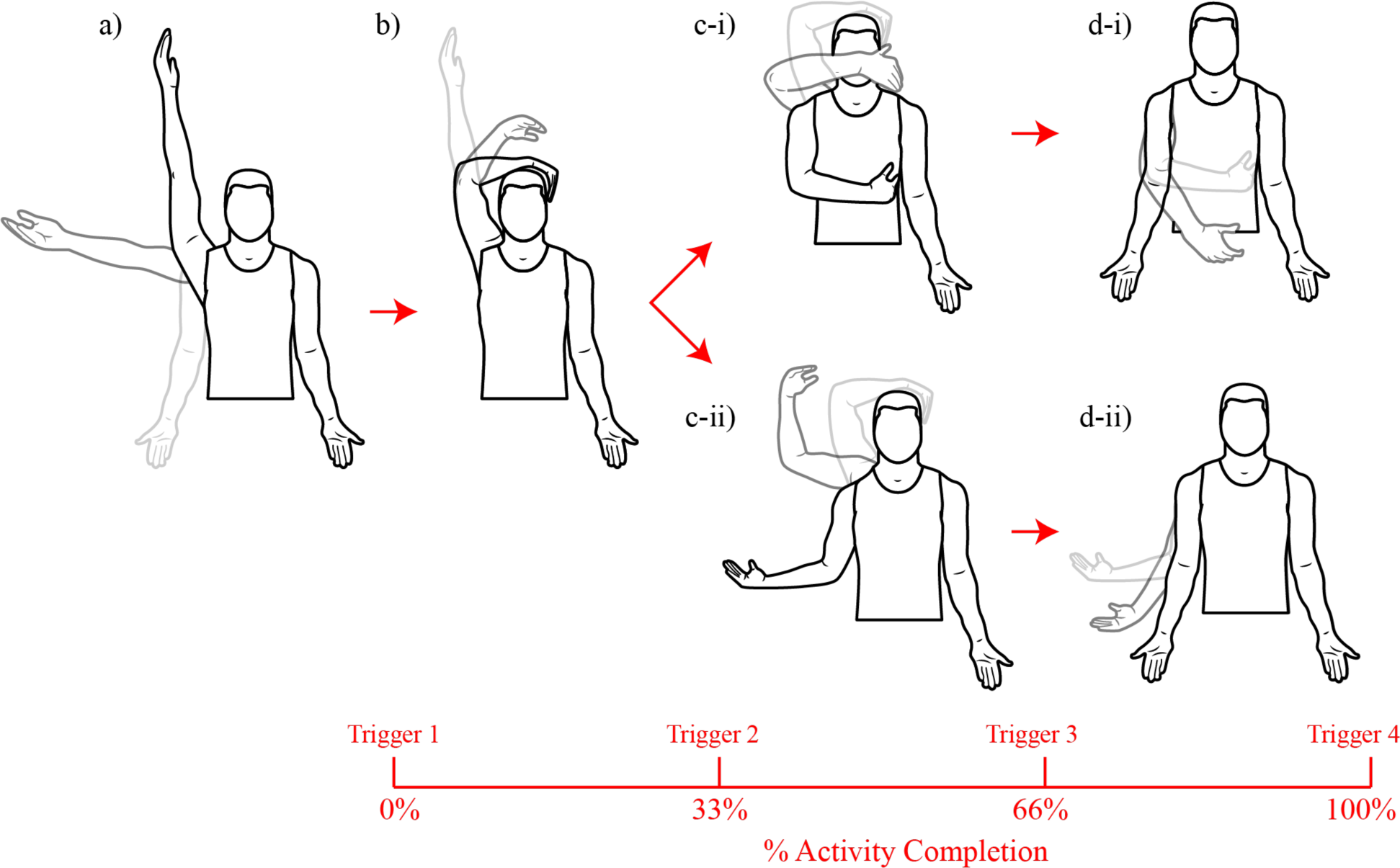
Illustration of the movements performed by participants while wearing the kirigami shoulder patch. a) The participant begins with their arm to the side and ends with their arm above their head. b) The participant begins with their arm above their head and ends with their forearm resting on their head. After completing movements a-b, participants were instructed to lower their arms in one of two paths. c-i) The participant begins with their forearm resting on their head and moves their arm in front of them in the sagittal plane into internal rotation, followed by d-i) the participant begins with their arm in internal rotation and moves it back into neutral position. Alternatively, c-ii) The participant begins with their forearm resting on their head and moves their arm to the side, in the coronal plane, into external rotation, followed by d-ii) the participant begins with their arm in external rotation and moves it back into a neutral position. Finally, four triggers were used to signal important time points in the movement. The timing of these triggers relative to the movements performed is shown at the bottom of the figure.

### Data Analysis

When collecting data, the participants were asked to rest for one second between each movement phase, allowing a study team member to press a trigger button. This trigger allowed the team members to identify the key start and end times of each movement phase. There was a total of four triggers: 1) when the participant’s arm is straight up above their head, 2) when the participant’s elbow is at a right angle with their forearm resting on their head, 3) when the participant’s arm is in internal or external rotation, and 4) when the participant’s arm is back to their side in the anatomical position. The timing of these triggers is visually represented in **Figure 2**.

The baseline measurement of each strain gauge differed slightly due to several factors, including different participant body shapes and different shoulder circumferences. These effects were mitigated by zeroing the strain gauge signals, as participants held a neutral position for several seconds at the beginning of each trial. All resultant data was therefore measured as the change in strain voltage compared to this baseline. The data was filtered with MATLAB’s low-pass filter with a 0.001 passband frequency, and segmented at each trigger. For the analysis, the timing of each movement phase was normalized to ease comparisons across each testing condition. This temporal normalization was accomplished by linearly interpolating the data and then downsampling to 101 data samples. Each segment was then stitched back together before running the analysis, such that the first movement segment, ranging from 0 to 33% of the total movement, represented the participant moving their arm from straight above their head to their forearm resting on their head. The second movement segment, ranging from 33-66% of the total movement, represented the participant moving into either internal or external rotation. The last movement segment, ranging from 66-100% of the total movement, represented the participant moving from internal or external rotation back to a neutral position with their arm back to their side. This data for each of the four strain gauges was then averaged across all five trials for internal or external rotation, and then averaged across all participants, resulting in eight time-series data sets. The data analysis was executed via a custom MATLAB script (v2023a, Mathworks Inc., Natick, MA).

### Statistical Analysis

We evaluated differences in strain voltage change between the internal rotation and external rotation movement patterns using one-dimensional statistical parametric mapping (SPM)^19^. This analytical approach allowed us to examine the ability of the *kirigami* patch to detect differences between the two shoulder rotation movements for each of the four strain gauges, and to determine when differences occurred across the three movement segments.

SPM was executed using custom-written Python code in Jupyter Notebook using Python version 3.11, and the open-source spm1d Python package library (www.spm1d.org)^19^. For each strain gauge, the five internal or external rotation trials were evaluated for each participant using SPM to detect intra-subject differences between the internal and external rotation conditions, followed by a group analysis comparing the two movement conditions with SPM using averaged data sets (one internal rotation and the other external rotation) for each strain gauge and participant. In all applications of SPM, the critical threshold was calculated based on a significance level of α = 0.05. The points of statistical significance between internal and external rotation for each strain gauge were determined using a two-tailed paired t-test, resulting in four statistical curves (SPM{t}).

## RESULTS

The movement patterns for each strain gauge of the *kirigami* shoulder patch were consistent within each participant, as depicted for a representative participant completing five movements resulting in internal rotation or external rotation in **Figure 3**. This data shows that each of the five trials for each strain gauge within each activity appears similar to the others. This data also shows visual differences in some strain gauges between internal and external shoulder rotation.

**Figure 3:**
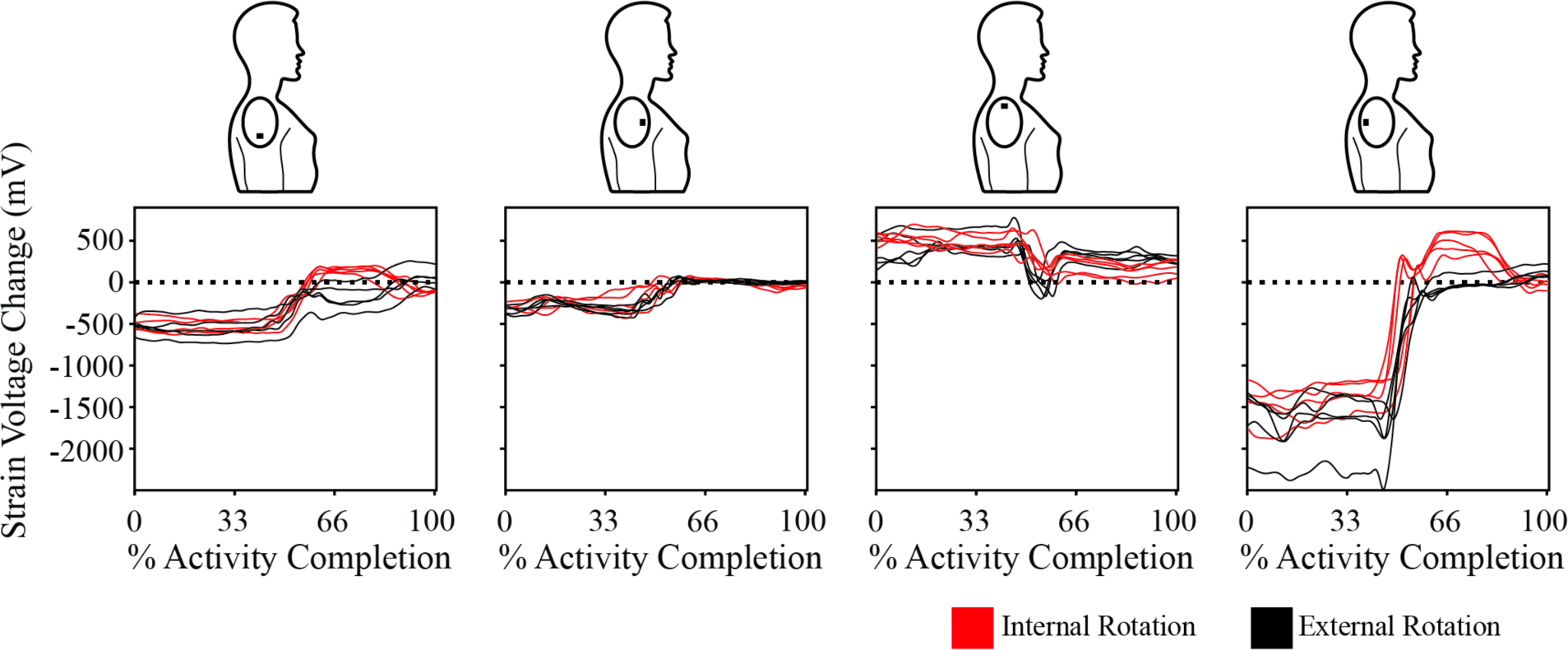
Representative data from one participant performing five trials resulting in internal rotation (red traces) or external rotation (black traces). The displayed data has been filtered and normalized to 100% of the activity movement. Each column shows resultant data from each of the four strain gauges, with the visible representation of the strain gauge location above each plot.

Intra-subject differences in movement patterns between internal and external shoulder rotation can be further examined using SPM. Intra-subject SPM models were performed on the movement patterns of all 17 participants, with the corresponding intra-subject results (separated by the three movement phases) shown in **Table 1**. In total, 14/17 individuals showed at least one significant difference between the two conditions. The most common movement phase where a significant difference was observed was between 33-66% of the movement when the arm is lowered into internal or external rotation.

**Table 1:**
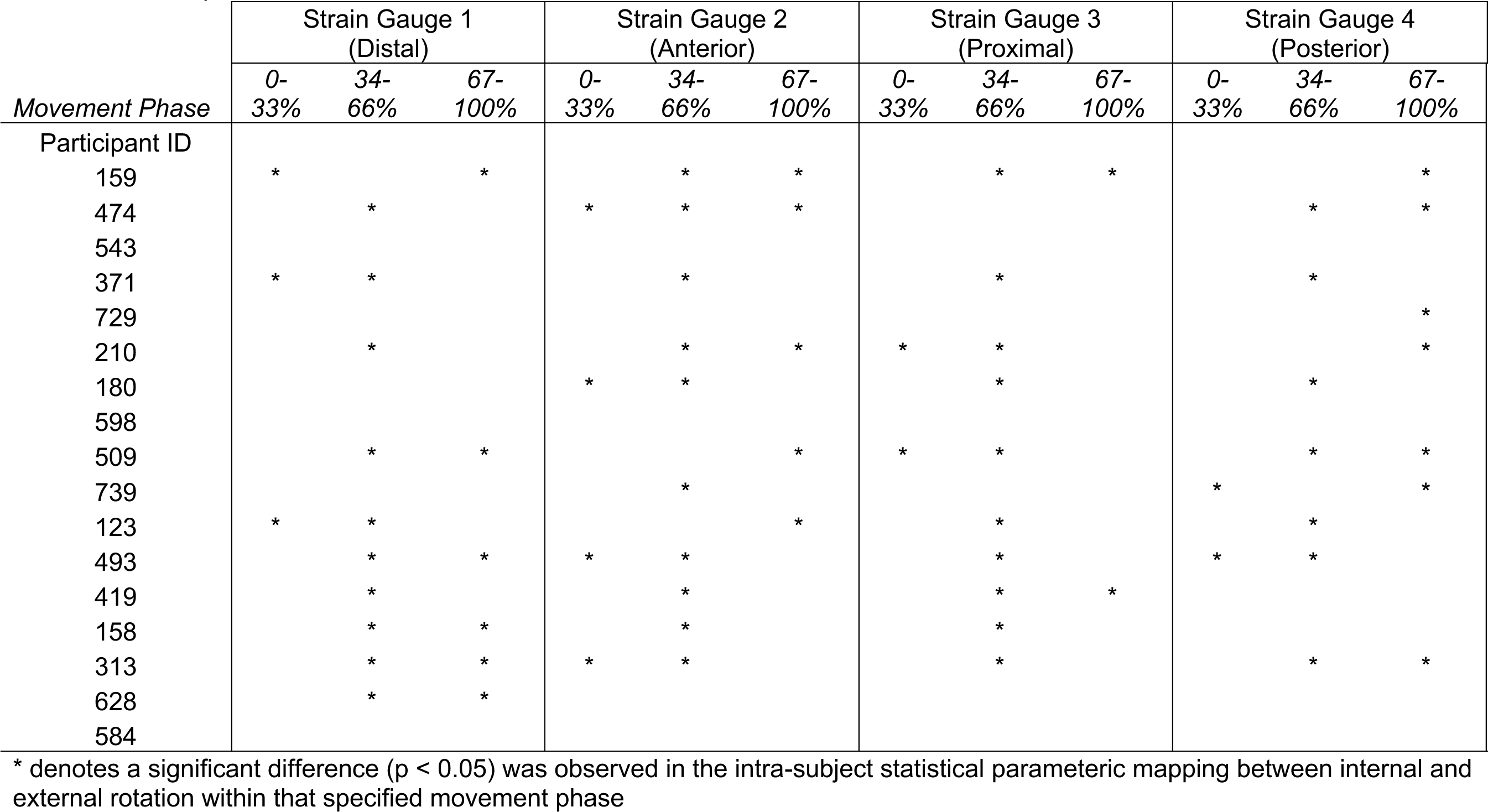
Summary of one-dimensional intra-subject statistical parametric mapping results for all 17 participants, striated by the three movement phases.

We extended our SPM approaches to look at movement differences between internal and external rotation movements across the entire group of 17 individuals. The group results are depicted in **Figure 4**. This approach yielded several significant findings. Strain gauge 4, located posterior to the acromion, exhibited the largest temporal differences between internal and external shoulder rotation and was significant between 31-41% and 51-90% of the movement (p = 0.047). Strain gauge 1, located distal to the acromion, was significant between 45-57% and 65-80% of the movement (p = 0.015). Finally, strain gauge 2, located anterior to the acromion, statistically differed between the internal and external shoulder rotation movements for a brief time between 51-54% and 80-87% (p = 0.039). Strain gauge 3, located proximal to the acromion, did not exhibit statistically significant temporal differences between internal and external shoulder rotation.

**Figure 4:**
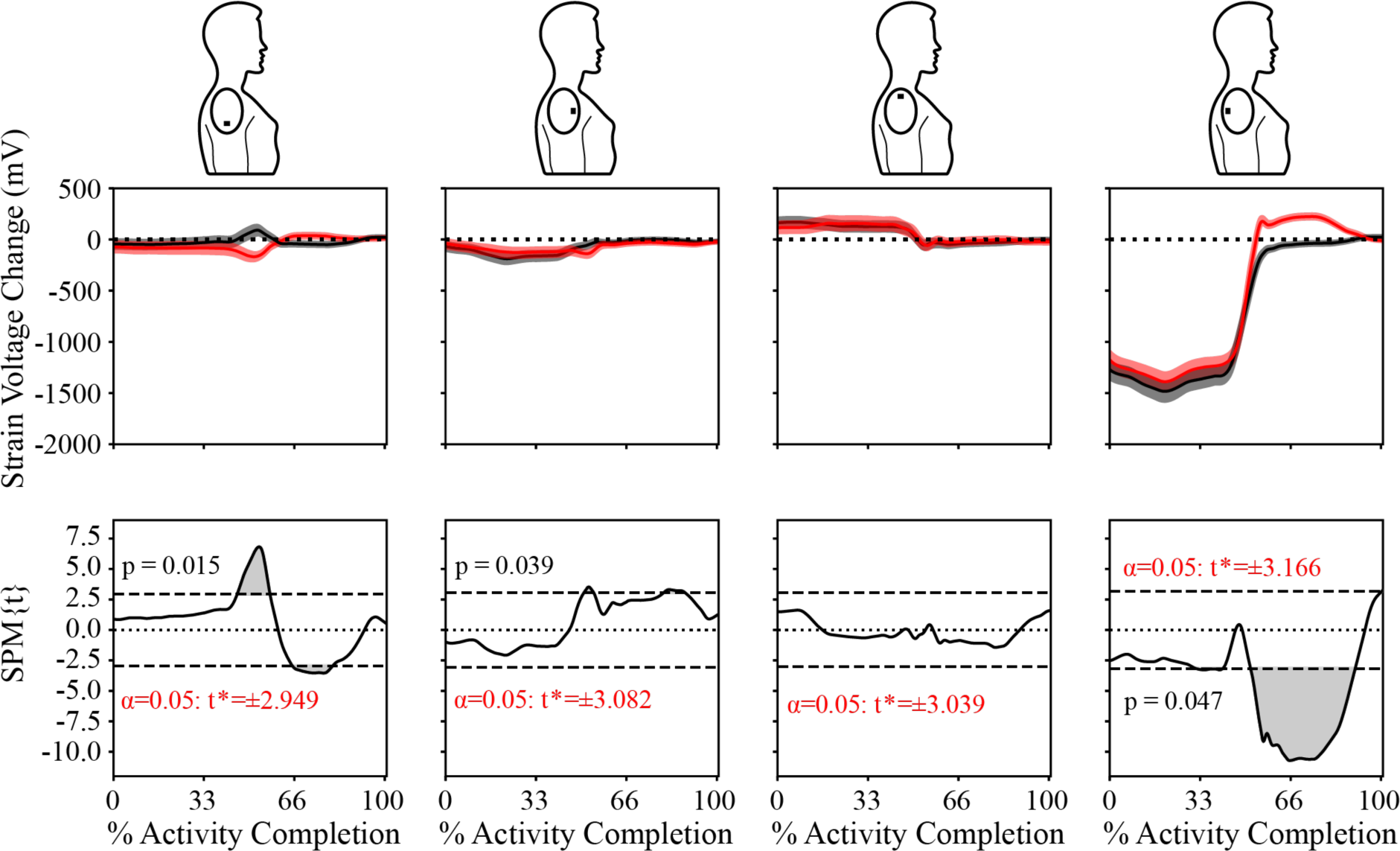
Data averaged across all 17 participants, with the top row showing the averaged times-series data for each of the four strain gauges after it has been filtered and normalized to 100% of activity movement. The mean of the five trials for external rotation is the bold black line with the standard error of the five trials shown in the gray shaded area. The mean of the five trials for internal rotation is the bold red line, with the standard error of the five trials for internal rotation in the pink shaded area. The bottom row displays the results from a t-test using statistical parametric mapping for each of the four strain gauges for the entire group, where the gray shaded area represents the area in which there is a statistical difference between internal and external rotation.

## DISCUSSION

The current study presents a novel investigation into the detection of humeral rotation using *kirigami* wearable sensor that conforms to the shape of the shoulder. The prescribed motions recorded here were particularly relevant to the study of Codman’s paradox, a phenomenon at the shoulder that allows for three degrees of rotation about the shoulder using two planar movements. The *kirigami* shoulder sensor, with its four on-board strain gauges, detected distinct differences in the movement pattern of participants who performed prescribed movements that resulted in either internal or external shoulder rotation. These results suggest wearable sensors that conform to the skin overlaying the shoulder can detect differences in skin deformation as the underlying humerus rotates into internal or external rotation.

The strain gauges most beneficial in detecting differences between internal and external rotation were the ones placed distal or posterior to the acromion. These locations are likely at a physiological advantage for detecting humeral rotation. The distal strain gauge is located over the humeral head, while the posterior strain gauge covers tightly linked fascial tissues overlying the posterior shoulder muscles. The posterior strain gauge exhibited a more significant signal change as the shoulder moved into internal than external rotation (between 67-100 percent of the movement). This would correspond with the tightening of the posterior capsule in response to internal rotation^20^.

Interestingly, the strain gauge covering the posterior shoulder was more sensitive to movement than the strain gauge over the anterior shoulder. These findings may indicate the posterior shoulder tissues are stiffer than the anterior tissues, as previously shown in throwing athletes^21^. The least beneficial strain gauge for detecting humeral rotation may be the one placed proximal to the acromion, the skin sensor farthest from the humeral head. While intra-subject differences for the proximal strain gauge were detected, the averaged group data did not show a difference. It is likely that the exact timing of when this difference was detected varied from participant to participant, and there was not a consistent phase of the movement that could therefore reach significance. The proximal strain gauge location may be more beneficial for detecting scapular movements, particularly elevation and upward rotation of the scapula. Unfortunately, the evaluated movement segments occurred after the arm was raised above the head, and therefore we likely did not capture any periods of elevation or upward rotation of the scapula within our three movement phases. Future work is therefore required to determine the effectiveness of the proximal strain gauge for measuring scapular movements.

The current study deploys SPM to evaluate temporal differences in shoulder kinematics. This analytic approach has been increasingly deployed over the past decade, allowing for type I and type II errors to be controlled at the time series level^22^. These approaches require a well-defined movement pattern, and thus, prior work has focused mainly on lower extremity applications given the cyclical gait pattern. There has been limited use of SPM analyses to measure shoulder kinematics before this study^23–27^. While this SPM approach works well for comparing tightly controlled shoulder movements like in the current study, this may not always be the case with shoulder movements, given the vast workspace in which to move and generate force with the upper extremity^28^.

Ultimately, this study aimed to use a new wearable technology to recognize shoulder movements, specifically internal and external shoulder rotation. The concepts of at-home monitoring and wearable sensors are not novel, as individuals have used glucose monitors and smartwatches to monitor blood sugar and activity levels. Despite the wide range of data collected and activities monitored with existing wearables, there is a long way to go before we achieve ubiquitous health monitoring, particularly for the monitoring of shoulder kinematics. Future work into *kirigami* shoulder sensors is required to address issues of cost, deployment, and clinical utility in diagnosing and managing shoulder pathologies.

The work described here has several limitations. First, the current study is a convenience sample of 17 healthy individuals, and it’s likely the evaluated movement patterns would differ in individuals experiencing shoulder pain or injury. Second, only a single size of the *kirigami* shoulder patch was used in the study. Given the differences in shoulder circumferences in a population, some of the variability measured could be due to the strain gauges sitting over slightly different musculoskeletal structures depending on whether the participant had a smaller or larger shoulder circumference. Third, the timing of each movement phase differed between participants. To ensure we were comparing the same movement phases, our research team had to normalize the timing of each movement segment in our analysis. Fourth, the current study relies on strain gauges measuring skin deformations around the shoulder as a proxy measure of shoulder kinematics. Shoulder kinematics using more established methods like motion capture or IMUs was not performed simultaneously due to financial and time constraints, and we cannot directly relate the strain gauge signals to an exact shoulder angle. We attempted to control this issue by using a well-defined shoulder movement pattern. The data in **Figure 3** shows consistency among all five trials for each strain gauge during each task, suggesting the participant performed the task similarly each time. Finally, our use of SPM provides an accurate method for detecting differences in signals with similar patterns but different amplitudes. Future research is needed to extend these methods to analyze other shoulder movement patterns.

### Perspective

The current study introduced the use of *kirigami*-inspired wearable sensors to quantify differences in internal and external rotation of the shoulder. These new wearable sensors that can adhere and conform to the shoulder could provide meaningful feedback to clinical professionals and researchers to better understand shoulder kinematics outside a laboratory environment. We found that specific strain gauge locations, particularly distal and posterior to the acromion, were most sensitive to detecting kinematic differences between internal and external shoulder rotation as participants completed movements consistent with Codman’s paradox. Expanded use of these sensor platforms in the future may better inform clinical decision making over more traditional approaches for measuring shoulder kinematics (e.g., marker-based motion capture), given their lower cost and enhanced portability.

## Data Availability

All data produced in the present study are available upon reasonable request to the authors

## Acknowledgments

This work was financially supported by the U.S. National Science Foundation (Graduate research fellowship awarded to Amani Alkayyali), the University of Michigan College of Engineering Seeding To Accelerate Research Themes (START) program (Max Shtein and Alanson Sample), and the University of Michigan Rackham Graduate School Graduate Student Research Grant (Amani Alkayyali).

## Conflict of Interests Statement

The authors have no conflicts of interest related to the work outlined in this manuscript.

## Notes

Funding statement: This work was financially supported by the U.S. National Science Foundation (Graduate research fellowship awarded to Amani Alkayyali), the University of Michigan College of Engineering Seeding to Accelerate Research Themes (START) program (Max Shtein and Alanson Sample), and the University of Michigan Rackham Graduate School Graduate Student Research Grant (Amani Alkayyali).

### Competing Interest Statement

The authors have declared no competing interest.

### Author Declarations

The ethics committee at Imperial College London (United Kingdom) gave ethical approval for this work.

